# An evaluation of reproducibility and errors in published sample size calculations performed using G*Power

**DOI:** 10.1101/2024.07.15.24310458

**Authors:** Robert T. Thibault, Emmanuel A. Zavalis, Mario Malički, Hugo Pedder

## Abstract

**Background:** Published studies in the life and health sciences often employ sample sizes that are too small to detect realistic effect sizes. This shortcoming increases the rate of false positives and false negatives, giving rise to a potentially misleading scientific record. To address this shortcoming, many researchers now use point-and-click software to run sample size calculations.

**Objective:** We aimed to (1) estimate how many published articles report using the G*Power sample size calculation software; (2) assess whether these calculations are reproducible and (3) error-free; and (4) assess how often these calculations use G*Power’s default option for mixed-design ANOVAs—which can be misleading and output sample sizes that are too small for a researcher’s intended purpose.

**Method:** We randomly sampled open access articles from PubMed Central published between 2017 and 2022 and used a coding form to manually assess 95 sample size calculations for reproducibility and errors.

**Results:** We estimate that more than 48,000 articles published between 2017 and 2022 and indexed in PubMed Central or PubMed report using G*Power (i.e., 0.65% [95% CI: 0.62% - 0.67%] of articles). We could reproduce 2% (2/95) of the sample size calculations without making any assumptions, and likely reproduce another 28% (27/95) after making assumptions. Many calculations were not reported transparently enough to assess whether an error was present (75%; 71/95) or whether the sample size calculation was for a statistical test that appeared in the results section of the publication (48%; 46/95). Few articles that performed a calculation for a mixed-design ANOVA unambiguously selected the non-default option (8%; 3/36).

**Conclusion:** Published sample size calculations that use G*Power are not transparently reported and may not be well-informed. Given the popularity of software packages like G*Power, they present an intervention point to increase the prevalence of informative sample size calculations.

## Introduction

Power calculations are used to identify the sample size required to test a specific hypothesis with predetermined false positive and false negative error rates, given various assumptions. Despite the substantial attention that low statistical power has received in the life and health sciences over the past decades, this issue continues to undermine research efforts (e.g., Border et al., 2019; Button et al., 2013; Lakens, 2022; Lamberink et al., 2018, Marek et al., 2022; Smaldino & McElreath, 2016).

A variety of reporting guidelines stress the use of appropriate sample sizes. These include the CONSORT statement (Moher et al., 2012: item 7a), STROBE statement (von Elm et al., 2008: item 10), ARRIVE guidelines (Percie du Sert et al., 2020: item 2b), the transparency checklist for social and behavioral research (Aczel et al., 2020: item 4 of 12), discipline specific reporting statements (e.g., Ros et al., 2020: item 1b) and journal reporting checklists (e.g., Nature Publishing Group, 2019). Other efforts include preregistration templates (OSF, 2016), the Experimental Design Assistant (Percie Du Sert et al., 2017), and Research Ethics Committees (Papalouka et al., 2023) that ask researchers to justify their chosen sample size. At least two studies analyzed the impacts of these policies and found that, after journals requested sample size justifications, more articles in those journals commented on sample size, but formal sample size calculations remained uncommon (e.g., a power or precision calculation) (Carter et al., 2017; The NPQIP Collaborative Group, 2019).

Of the published studies that do use power calculations, many of the calculations may be irreproducible or contain errors (e.g., Charles et al., 2009; Clark et al., 2013; Rutterford et al., 2015). G*Power is a widely used sample size software for non-statisticians. However, if the user does not understand the underlying statistics, the ease of using its graphical user interface can lead to erroneous calculations. For example, if users are unaware of the default option for mixed-design ANOVAs, they may accidentally account for the correlations between the repeated measure a second time, and in turn substantially—but erroneously—increase power (see Figure 1; this issue is also explained in further detail by Kieslich, 2020; Lakens, 2013; and Thibault & Pedder, 2022). The present study set out to estimate the prevalence of irreproducible or erroneous power calculations performed in published articles using G*Power.

**Figure 1.**
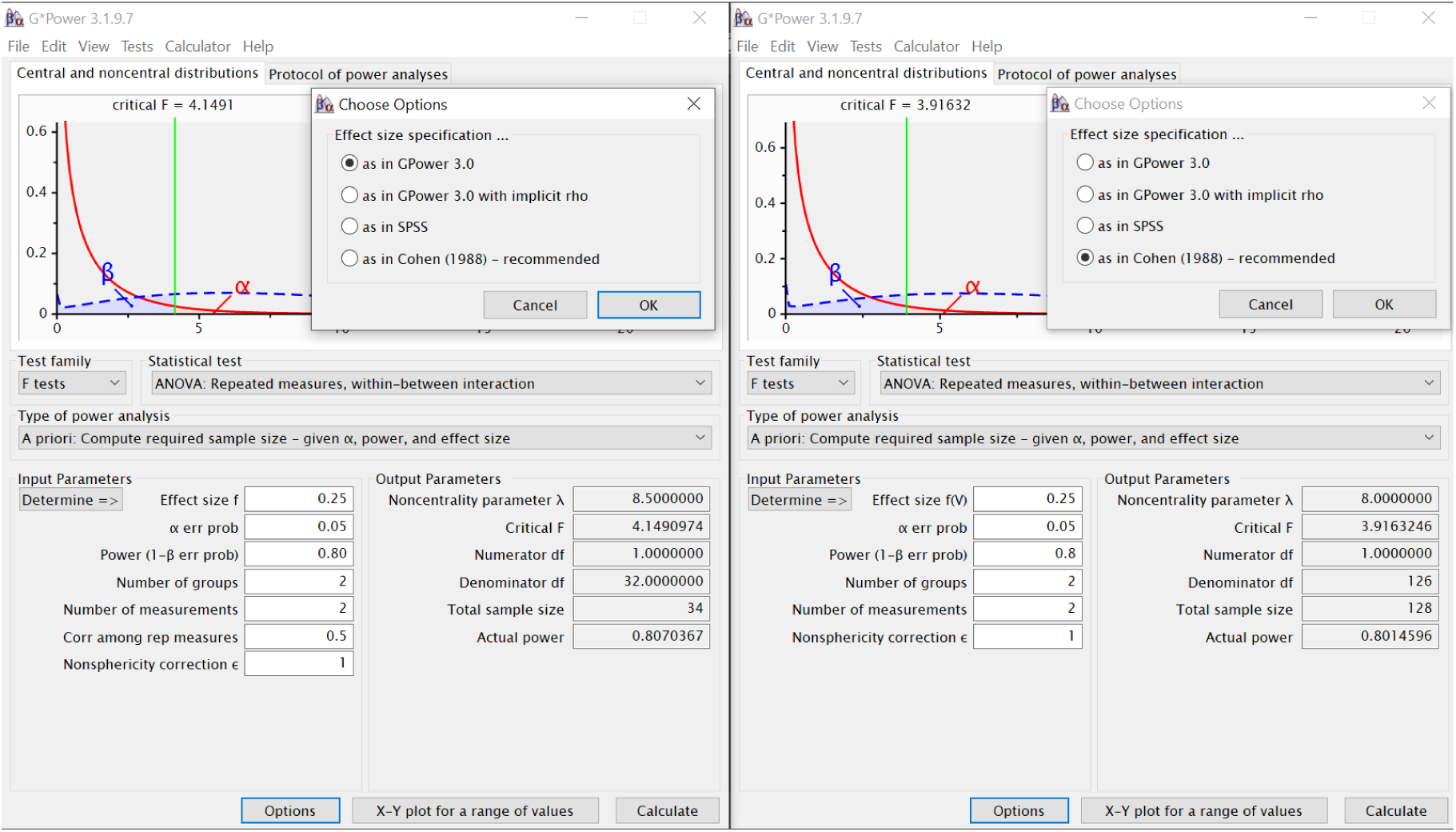
Depiction of the G*Power options for a mixed-design ANOVA. Depending on the option selected, the software calculates a sample size of 34 or 128. Note, the G*Power software uses the term “ANOVA: Repeated measures” for what is commonly called a mixed-design ANOVA: an analysis with one within-subjects factor and one between-subjects factor (rather than only within-subjects factors). The *Statistical test* drop-down menu includes three choices for “ANOVA: Repeated measures”: between factors, within factors, and within-between interaction. The issue we describe is apparent for within-between interactions and within factors. In our experience, most sample size calculations for mixed-design ANOVAs are interested in within-between interactions.

### Box 1.

A word on terminology

The introduction of this article uses the term **power calculation**. This term comes with a degree of ambiguity and some might consider it a misnomer. Power calculations are generally used to indicate an equation that solves for sample size, based on a *p*-value (alpha), power (1-beta), and an effect size. However, the equation can be rearranged to solve for any of these four variables, and many people would still call it a power calculation.

In this paper we use the term **sample size calculation** when a power calculation solves for sample size. Some people call this an *a priori* power calculation, because it should be done before running a study—but the misnomer remains, as the equation is not solving for power.

If the equation is rearranged to solve for power, and the exact *p*-value discovered in a study is used as an input to the calculation, this is a ***post-hoc* power calculation**. *Post-hoc* power calculations are recommended against and can be misleading (Goodman and Berlin, 1994).

Rearranging the equation to solve for effect size is often called a **sensitivity analysis** and is applicable when researchers are working with a fixed sample size (e.g., when analyzing a dataset that already exists).

**Precision calculations** are an alternative to power calculations. Instead of calculating a sample size to show whether or not an effect of a certain magnitude exists, they calculate the sample size needed to estimate the effect size with a certain margin of error (e.g., for a 95% confidence interval). For example, we could calculate the sample size needed to show that a diet intervention leads to a reduction of at least 5 kilograms (power calculation), or we could calculate the sample size needed to estimate the degree of weight loss within ±3 kilograms regardless of the point estimate (a precision calculation). While precision calculations are preferable in many situations—we’ve used one for this study—this paper focuses on power calculations as they are more common.

**Mixed-design ANOVAs** compare the means between independent groups on a repeated measure. In other words, there is one *between-subjects* factor (the group) and one *within-subjects* factor (e.g., measurement at multiple time points).

### Study objectives

This study is primarily descriptive. The objectives are to:

1. Estimate the number of published articles that use G*Power to perform a sample size calculation.
2. Assess the reproducibility of sample size calculations that use G*Power in published articles.
3. Estimate the prevalence of error-free sample size calculations that use G*Power in published articles.
4. Estimate the prevalence of using G*Power’s default option for mixed-design ANOVAs, which can be misleading (see Figure 1).

## Methods

### Sample Size

We aimed to estimate the prevalence of reproducible sample size calculations and error-free sample size calculations that use G*Power. We performed a precision analysis to inform our sample size (Rothman & Greenland, 2018) using Monte Carlo sampling for 95% confidence intervals (outlined in the protocol code available at osf.io/dsv4m). This analysis returns very similar sample sizes to the equation *n* = *(z*^2^ *\* p(1 — p))/ MOE*^2^, which is often used for precision analyses. This equation, however, relies on a normal approximation to the binomial distribution, which doesn’t hold true for small sample sizes or proportions near 0 or 1.

The majority of the questions on our coding form are binary and thus, we can use the same precision calculation for all these variables^1^. We used a 95% confidence interval with a maximum width of 20% between the lower bound of the confidence interval to the upper bound, and corresponding to the most conservative expected proportion of 0.50. This requires a minimum sample size of 95 articles.

We ran an additional precision analysis for Objective 4, which asks whether power calculations for mixed-design ANOVAs clearly select the appropriate option related to their analysis (see Figure 1). This question applies to only a subset of the articles we sampled. We expected that about 20% of relevant articles would clearly select the appropriate option related to their analysis^2^. This expected proportion of 0.20 requires a sample size of 60 articles. With a sample size of 60 articles, the 95% CI could have a width of up to 26% (i.e., ± 13%, if the sampled proportion turns out to be 0.50), which we deem acceptable for our purposes.

### Sample

We restricted our study to articles published after 2017 to assess recent practices. To identify published articles that used G*Power to run a power calculation we used the search query *(GPower OR “G Power”) AND (“2017/01/01”[Publication Date] : “3000”[Publication Date])* in PubMed Central (PMC), on 31 May 2022^3^. Note, searches in PMC convert the characters “*” and “-” to a space (when they are in the middle of the word that is within quotation marks). This makes the terms *“G Power”, “G*Power”, and “G-Power”* equivalent in the query. We searched PMC because, unlike PubMed and most other databases, PMC only indexes open access articles and search queries scan the full text article rather than only the title, abstract, and keywords.

We downloaded the full list of 22,188 PMCIDs returned from our search query. We then randomly sampled 500 PMCIDs using the *sample_n()* function in R with seed number 1313. We sequentially selected PMCIDs from that list until we reached the target sample size of 95 articles that report performing a sample size calculation using G*Power. We also counted the number of articles that perform power calculations using G*Power that solved for variables other than sample size (e.g., for power or effect size). However, we did not code these calculations for reproducibility or errors. We excluded articles that discuss G*Power without using it to perform a power calculation, as well as tutorials on how to use G*Power.

We sampled additional articles that contained G*Power calculations that solved for sample size specifically for a mixed-design ANOVA, until we reached 36^4^. Thus, we sampled more than 95 articles in total. However, we report all results in terms of the 95 articles, except for results concerning mixed-design ANOVAs.

### Data extraction

We created a coding form specifically designed for this project (available in Appendix A of the protocol: https://osf.io/xhfn8). This form includes three overarching sections. We first assessed whether the power calculation solved for sample size, power, effect size, or another variable. Second, we assessed whether the article transparently reported key variables of the power calculation—including alpha, power, sample size, effect size, and the statistical test. We also tried to reproduce each calculation using G*Power. Third, we assessed whether there were errors in the power calculations, including whether the sample size calculation matched the statistical analysis used in the results section of the article, which mixed-design ANOVA option was selected, and other visible errors (e.g., inputting a non-standardized effect size into a calculation that requires a standardized effect size, stating that Cohen’s *d* = 0.2 is a conventionally large effect size, claiming a two-tailed test when a one-tailed test was run). The coding form also included a free-text response for types of errors we did not prespecify. If an article had more than one sample size calculation we only extracted information from the primary sample size calculation, or the first-listed sample size calculation if none was specified as primary.

Two investigators (EAZ & RTT) independently coded each article and resolved coding differences through discussion. If necessary, an additional investigator arbitrated (HP). Investigators had the option to select that they did not have the statistical expertise to code an article. If they selected this option, a statistician from our team (HP) coded that article. Inter-rater agreement ranged from low to high depending on the specific item. Precise inter-rater agreement values are included in Supplementary Material C alongside explanations that may account for the low agreement on select items.

### Statistical Analysis

We estimate 95% CIs and point estimates for all proportions and numbers using Monte Carlo sampling, as outlined in the analysis code. We round several numbers in the results section to avoid implying greater precision than we have (although we maintained the full values throughout the analysis steps).

## Results

### Objective 1: How many articles use G*Power for a sample size calculation?

Our query in PMC revealed that 22,188 of the 3,285,893 articles published between 1 Jan 2017 and 31 May 2022 and indexed in PMC mentioned G*Power. To achieve our target sample size of 95 articles, we sampled 147 articles. 141 (96%) of these articles reported using G*Power for a power calculation. Thus, roughly 21,000 (22,188 * 0.96) articles indexed in PMC between Jan 2017 and May 2022 report using G*Power for a power calculation. This equates to about 48,000 articles in PubMed^5^ (Supplementary Table 1).

We arrive at similar estimate based on the number of citations to the two main published articles from the G*Power team (Faul et al., 2007; Faul et al., 2009). These articles have about 34,000 and 60,000 citations respectively on Google Scholar, and 16,000 and 35,000 citations on Web of Science (as of 6 Nov 2023).

In our sample, only 67% (95/141) of the G*Power calculations solved for sample size (Table 1). This corresponds to approximately 32,000 publications in PubMed (Supplementary Table 1). We therefore estimate that 1 out of every 154 articles on PMC report using G*Power for a power calculation (i.e., 0.65% [95% CI: 0.62% - 0.67%] of articles), and about 1 out of every 229 articles on PMC report using G*Power for a sample size calculation (i.e., 0.44% [95% CI: 0.38 - 0.49%]).

**Table 1.**
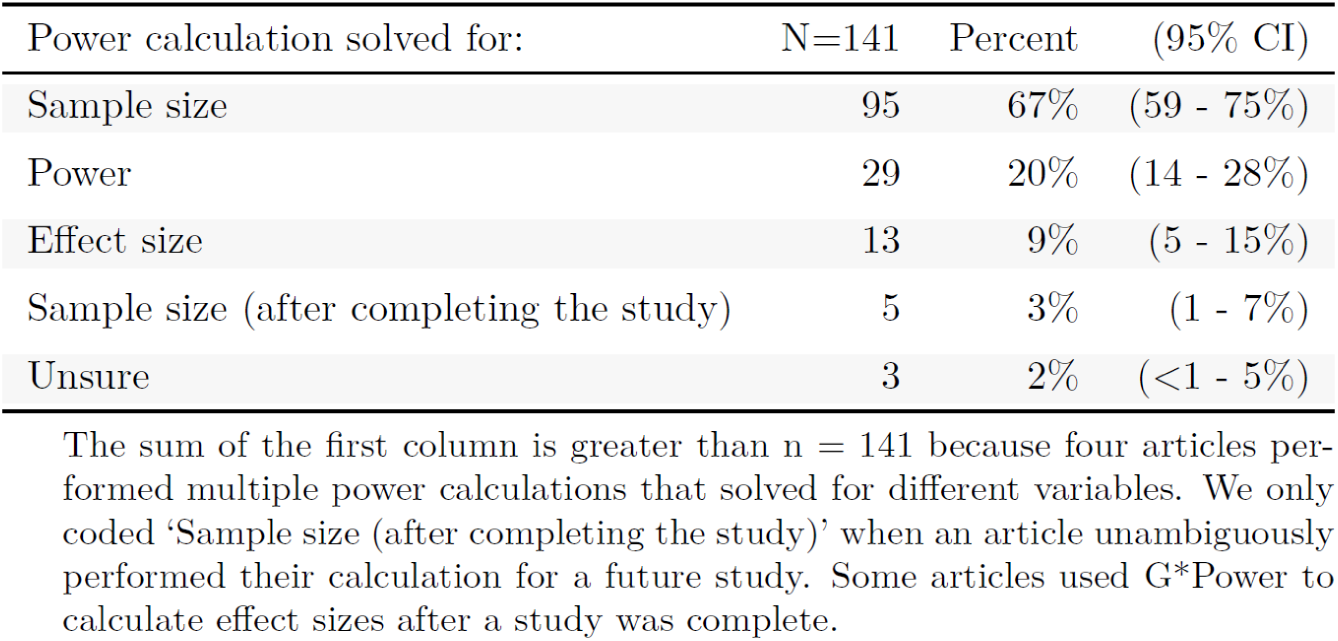
Types of power calculations.

| Power calculation solved for: | N=141 | Percent | (95% CI) |
| --- | --- | --- | --- |
| Sample size | 95 | 67% | (59 - 75%) |
| Power | 29 | 20% | (14 - 28%) |
| Effect size | 13 | 9% | (5 - 15%) |
| Sample size (after completing the study) | 5 | 3% | (1 - 7%) |
| Unsure | 3 | 2% | (<1 - 5%) |

The sum of the first column is greater than $n = 141$ because four articles performed multiple power calculations that solved for different variables. We only coded 'Sample size (after completing the study)' when an article unambiguously performed their calculation for a future study. Some articles used G\*Power to calculate effect sizes after a study was complete.

### Objective 2. How reproducible are the sample size calculations?

Based only on the information reported in the article, we were able to reproduce 2% (2/95) of the sample size calculations (i.e. the authors provided all parameters needed to be entered into G*Power). After making assumptions (e.g., alpha = 0.05, power = 80%, independent vs. paired-sample, number of tails, number of factor levels)^6^ we were likely^7^ able to reproduce an additional 28% (27/95) of the sample size calculations. We could not reproduce the remaining 70% (66/95)^8^. We also calculated the percentage of study participants who took part in studies with sample size calculations that we could reproduce. We found that 27% (3,083/11,469) of participants took part in a study with sample size calculations that we could reproduce or likely reproduce.

Alpha and power were almost always reported, and they rarely deviated from commonly used values (α = 0.05, power = 0.80 or 0.95) (Table 2). The effect size type and statistical test used were often not reported (Table 2). Only 14% (13/95) of published sample size calculations reported all of alpha, power, effect size type, effect size value, sample size, and statistical test^9^.

**Table 2.**
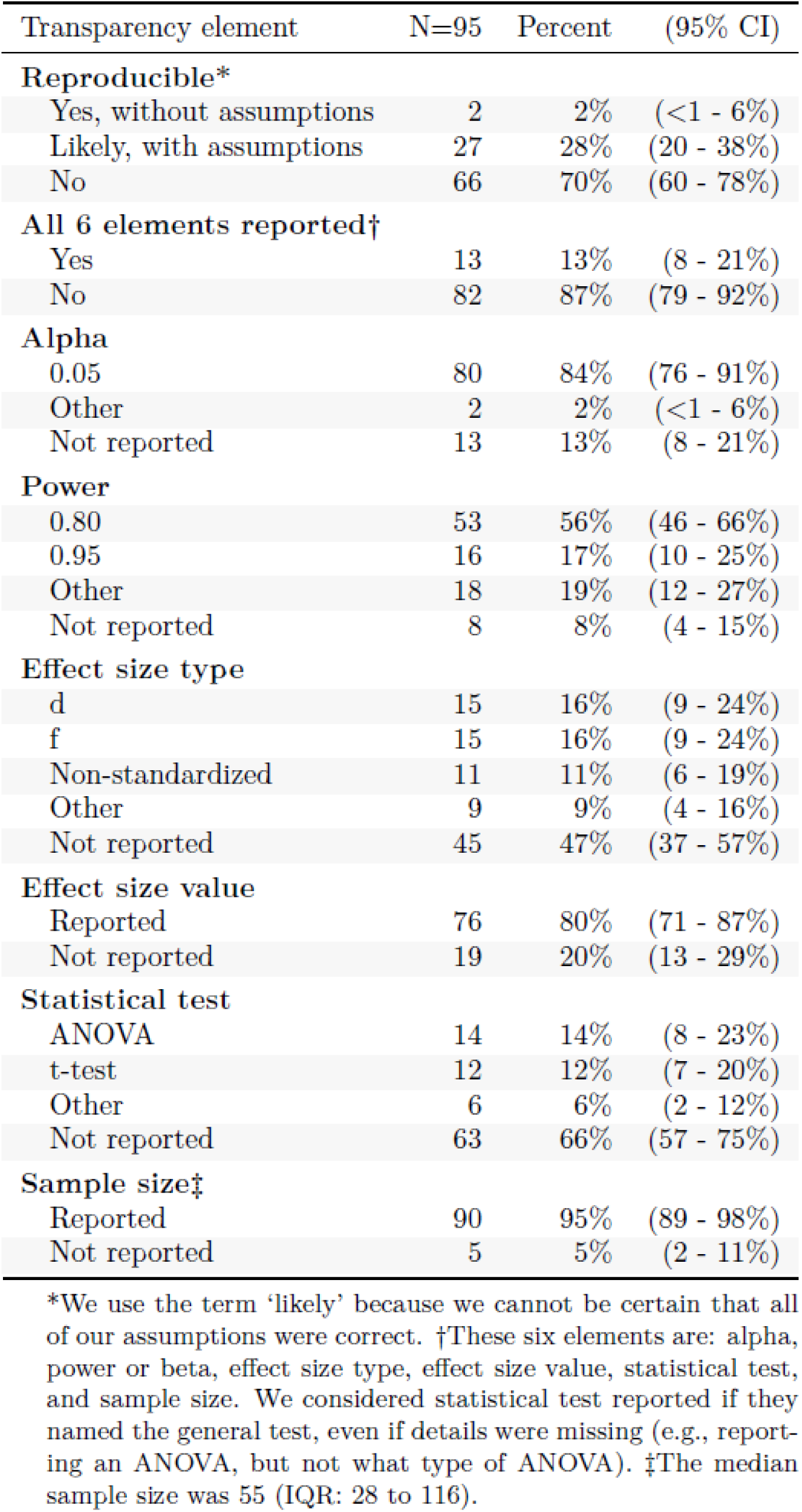
Reproducibility of sample size calculations performed using G*Power.

Details about the types of articles we analyzed are presented in Table 3, as well as in the *journal* and *publisher*^10^ columns of our open data. The median sample size reported in the sample size calculations was 55 (IQR: 28 to 116).

**Table 3.** Article characteristics.

| Article characteristic | N=95 | Percent |
| --- | --- | --- |
| **Unit of study** |  |  |
| Human | 85 | 89% |
| Non-human animal | 10 | 11% |
| **Year of publication** |  |  |
| 2017 | 4 | 4% |
| 2018 | 8 | 8% |
| 2019 | 20 | 21% |
| 2020 | 29 | 31% |
| 2021 | 22 | 23% |
| 2022 | 12 | 13% |
| **Protocol article** |  |  |
| Yes | 8 | 8% |
| No | 87 | 92% |
| **Multiple sample size calculations** |  |  |
| Yes | 5 | 5% |
| No | 90 | 95% |
| **Publisher** |  |  |
| BMC | 18 | 19% |
| MDPI | 14 | 15% |
| Frontiers Media SA | 7 | 7% |
| PLOS | 6 | 6% |
| Springer | 5 | 5% |
| Wiley | 5 | 5% |
| Nature Portfolio | 3 | 3% |
| Other | 37 | 39% |

The median journal impact factor was 2.7 (range: 1.4 to 11). 18 articles were published in journals that did not have an impact factor. Year 2023 contains fewer articles than the preceeding years because we only sampled until May 31, rather than the entire year.

### Objective 3. Are the sample size calculations error-free and the effect sizes justified?

For 48% (46/95) of the articles, we were unsure whether the statistical test used in the sample size calculation matched a statistical test used in the article^11^. For 30% (29/95) of articles, we found a statistical test in the results section that broadly matched the one mentioned in the sample size calculation. For 13% (13/95) of articles we could not find a matching statistical test. (Table 4).

**Table 4.** Quality of the sample size calculations performed using G*Power.

| Quality measure | N=95 | Percent | (95% CI) |
| --- | --- | --- | --- |
| **Analysis match in results section** |  |  |  |
| Yes | 29 | 30% | (22 - 40%) |
| No | 13 | 13% | (8 - 21%) |
| Unsure | 46 | 48% | (38 - 58%) |
| NA (protocol) | 7 | 7% | (3 - 13%) |
| **Error** |  |  |  |
| Yes | 10 | 10% | (5 - 17%) |
| No | 14 | 14% | (8 - 23%) |
| Unsure | 71 | 75% | (66 - 83%) |
| **Adjusted for multiple comparisons** |  |  |  |
| Yes | 0 | 0% | (NA) |
| No, and multiple analyses are performed | 71 | 75% | (66 - 83%) |
| No, but a single outcome was identified | 22 | 23% | (15 - 32%) |
| Unsure | 2 | 2% | (<1 - 6%) |
| **Justification for chosen effect size\*** |  |  |  |
| Previously published research | 25 | 26% | (18 - 35%) |
| Effect size conventions | 25 | 26% | (18 - 36%) |
| General reference to another study | 9 | 9% | (4 - 16%) |
| Pilot data | 5 | 5% | (2 - 10%) |
| Effect size of interest | 4 | 4% | (1 - 9%) |
| Other | 3 | 3% | (1 - 7%) |
| No justification reported | 31 | 33% | (24 - 42%) |

\*Some articles provided more than one justification for their chosen sample size, and thus the sum of the percentages is greater than 100%.

We were unsure whether 75% (71/95) of the articles had an error related to their sample size calculation. We deemed 14% (14/95) to be error-free, and 10% (10/95) to include an error (Table 4). Examples of errors include providing a sample size that is impossibly small for the effect size they reported using, and powering for a t-test when the primary outcome is binary, among others. Additional information is available in our open data in the *error_text* column.

None of the sample size calculations accounted for multiple comparisons. Although 76% of articles ran multiple statistical tests and did not specify a primary outcome or main statistical test (Table 4).

4% (4/95) of sample size calculation used an effect size of interest; 26% (25/95) referenced previously published effect sizes; 9% (9/95) made a general reference to another study to justify their sample size calculation, but not specifically to the previous study’s effect size. Another 26% (25/95) referenced conventions for sample size (e.g., selected a medium effect size of Cohen’s d = 0.5). Finally, 33% (31/95) did not provide a justification for the effect size they used (Table 4).

### Objective 4. How often do researchers select the non-default G*Power option for mixed-design ANOVAs?

8% (3/36) of sample size calculations that used a mixed-design ANOVA selected the non-default option (Table 5). Half (18/36) used the default option and 42% (15/36) were unclear. Although we could not reproduce the 15 calculations that were unclear, their sample sizes were often closer to what would be produced if the default option, as opposed to non-default option, was selected. In our dataset, articles that used the default option calculated sample sizes that were between 3 to 8 times smaller^12^ than the sample size they would have calculated if using the non-default option (which is generally the more correct option to choose: Lakens, 2013; Thibault & Pedder, 2022).

**Table 5.** Selection of default ANOVA option.

| Option used | N=36 | Percent | (95% CI) |
| --- | --- | --- | --- |
| Non-default option | 3 | 8% | (2 - 19%) |
| Default option | 18 | 50% | (34 - 66%) |
| Unsure | 15 | 42% | (26 - 58%) |

## Discussion

### Summary

We estimate that 48,000 articles indexed in PMC or PubMed report using G*Power for a power calculation from 2017-2022. About two-thirds of these calculations solve for sample size. Few publications transparently report the details of their sample size calculations, which left our team unable to reproduce 70% of the calculations, and made it difficult to assess whether the calculations contained errors. Very few articles selected the non-default option for mixed-design ANOVAs—which may have led researchers to use sample sizes far too small to answer their intended question.

### Limitations

Due to the limited reporting transparency in many of the sample size calculations we assessed, it was difficult to assert whether an error was present. It was also unclear at what time during the research life-cycle the sample size calculations were performed. Some calculations might have been performed during the manuscript writing phase in order to meet journal reporting requirements. Second, mega-journals (e.g., MPDI) account for a substantial portion of our sample. These journals may be systematically different from more traditional journals (Ioannidis et al., 2023) and could impact our results.

Third, we randomly sampled published articles and gave each article equal weighting. While this approach is common across meta-research studies, it treats each individual article as an equal output. Studies with more resources and more participants might have better reporting. For example, a preliminary analysis found articles in top medical journals had transparent sample size calculations (Thibault et al., 2023). Although, we were able to measure the rate of reproducible sample size calculation per participant, and found comparable values to when measuring per article (27% versus 30%, respectively). If we had the information to measure the rate of reproducible and error-free sample size calculations based on the amount of research funding spent, the rate of reproducible and error-free sample size calculations may be greater than what we report in the present manuscript.

#### Box 2.

**Cosplay power calculations**

50 years ago, the physicist Richard Feynman coined the term Cargo Cult Science to describe research that has the outward appearance of being scientific, but which is missing an essential element that makes it scientific—which he defined as bending over backwards to ensure we are not fooling ourselves (Feynman, 1974). The results from the present manuscript suggest that some power calculations could be described this way. We opt for the term “Cosplay”^13^ Power Calculations, due to the potential cultural insensitivity of the term Cargo Cult. 20% of calculations we assessed solved for power. Looking at the text for these articles (viewable in the *verbatim* column of the open data) reveals that many calculations were performed after study completion (i.e., they are *post-hoc* power calculations). Some authors also use these calculation to make a circular argument that their study had sufficient power, or didn’t have sufficient power, to find the effect size which they already found^14^.

A few articles ran a sample size calculation after completing their study. For example, to calculate the sample size required to reach significance given their findings (“…*due to a lack of power, we performed sample size calculations to estimate the sample size that would be required to be able to observe a significant interaction effect…We would have needed a sample size of 4208 for the insula”*—PMC6162305). This approach treats the size of an effect as irrelevant. Based on the direct relation between sample size and *p*-values, any effect size can become statistically significant when enough participants are sampled. Most sample size calculations we assessed (75%) did not include enough detail to identify whether an error was present, and in turn, whether or not the sample size calculation was meaningful. Taken together, a non-negligible portion of published articles include Cosplay Power Calculations—they are dressed up to fill a meaningful role, but are unable to deliver their key purpose.

### Smallest effect size of interest

Ideally, an effect size for a power calculation is chosen because it is meaningful (e.g., a smallest effect size of interest or Minimal Clinically Important Difference—MCID). Only 4% of calculations we reviewed provided such a justification. The other calculations either did not provide a justification (33%) or used shortcuts that fail to provide insight as to whether the effect size is meaningful in the context of the specific study (e.g., referencing another study; using conventions about whether the effect size is small, medium, or large). Notably, there is no broadly-accepted standard for how to determine what effect size to consider meaningful, and in turn use in a sample size calculation. Nonetheless, we feel that a researcher performing a sample size calculation should be able to provide some degree of justification for why their chosen effect size could be meaningful.

### Impact of poorly-informed sample size calculations

Many studies have already established that the published literature across many disciplines relies on sample sizes that are too small to detect realistic effect sizes, and that this practice leads to an untrustworthy knowledge base (e.g., Border et al., 2019; Button et al., 2013; Lakens, 2022; Marek et al., 2022; Smaldino & McElreath, 2016). Ideally, researchers who perform sample size calculation would be overcoming this issue. However, if errors exist in their sample size calculations, the calculation may simply mask that their sample sizes are too small to answer their question^15^. Whether the sample sizes are indeed too small can be determined by checking whether the effect size of interest falls within a confidence interval based on the observed data (Goodman and Berlin, 1994); yet this information is rarely interpreted for this purpose.

To put this issue in context, the median sample size from the calculations we assessed was 55. If we were planning to recruit 55 participants, divide them into a treatment and control group and then compare their outcomes (e.g., via an independent-sample t-test) we would be able to detect an effect size of Cohen’s *d* = 0.99 (with 95% power) and *d* = 0.77 (with 80% power)—see Supplementary Figures 1 and 2. For comparison sake, the height difference between men and women over the age of 20 in the United States is *d* = 1.01 (National Center for Health Statistics, 2021)^16^ and the mean effect size of the 20 most common pharmaceutical therapies is *d* = 0.58 (median *d* = 0.56) (Leucht et al., 2015). Thus, many studies in our sample used sample size calculations that were only powered to detect effects that are larger than some of the most widely used pharmaceuticals and as distinguishable as the height difference between men and women.

The issue is not that software packages like G*Power drive researchers to use small sample sizes; it’s that subpar sample size calculations can provide a false impression that a known problem (small sample size) is being addressed when it’s not. This issue becomes more concerning when scientific publications—which generally include multiple authors, multiple reviewers, and at least one editor—fail to address irreproducible and potentially erroneous sample size calculations.

### Recommendations

Our study touches on two issues in regard to sample size calculations: transparency and quality. Transparency is relatively easy to address compared to quality. To comprehensively report a power calculation that solves for sample size, authors must report at least six elements: alpha, power, effect size type, effect size value, statistical analysis, and sample size. For a sample size calculation to be reproducible without making assumptions, authors would also need to report specific details, such as how many tails were used and the exact type of test (e.g., paired sample t-test, two-tailed, with correlation coefficient of 0.5). All this information can be reported in a few sentences^17^, or by taking a snapshot of the G*Power screen and sharing that image. And yet, only 14% of sample size calculations reported all 6 of these elements within the text describing the sample size calculation. Sample size calculation software could be designed to output the details of the calculation, so they are reproducible. In parallel, journals that already require sample size calculations could also require that all parameters are reported.

To improve the quality of sample size calculations would take more concerted and persistent efforts. Ideally, every researcher would have a statistician available to help with sample size calculations. Given that this situation is unrealistic, it behooves us to create sample size calculation software that is tailored to researchers, who may not always have in-depth statistical skills. For example, removing default options can force users to consider their choices and avoid making uninformed default selections. This is particularly relevant given the impacts of the potentially misleading default option for mixed-design ANOVAs in G*Power.

## Conclusion

Sample size calculation software packages like G*Power may prove valuable to combat uninformative research due to small sample sizes. However, publications that report using this software generally provide too little information to assess whether their calculations were performed correctly. Moreover, it appears that the default setting for sample size calculations for mixed-design ANOVAs provides a substantial underestimation of the sample sizes required. As researchers in the life and health sciences try to improve the reproducibility and quality of their work, well-designed sample size calculation software could prove beneficial.

## Data availability

Data, data dictionaries, analysis scripts, and other materials related to this study are publicly available at https://osf.io/msz24/. The study protocol was registered on 31 May 2022 at https://doi.org/10.17605/OSF.IO/UJXHW. Discrepancies between this manuscript and the registered protocol are outlined in Supplementary Material A. The analysis script can be rerun by selecting “Reproducible Run” in the Code Ocean container available at https://doi.org/10.24433/CO.4349082.v1.

## Acknowledgements

We thank Steven Goodman and Marcus R. Munafò for feedback on the study protocol and insights provided through discussion about this project. We thank Steve Goodman and Tom E. Hardwicke for commenting on a draft manuscript.

## Contributions

Conceptualization: RTT, EAZ

Data curation: RTT

Formal analysis: RTT

Funding acquisition: RTT

Investigation: RTT, EAZ, HP

Methodology: RTT, EAZ, MM, HP

Project administration: RTT

Software: RTT

Supervision: RTT

Validation: RTT

Visualization: RTT

Writing - original draft: RTT

Writing - review & editing: RTT, EAZ, MM, HP

## Supplementary Material A. Deviations from the preregistered Protocol

We made the following deviations from the preregistered protocol.

- The final sample size for the mixed-design ANOVA analysis included 36 articles. We had originally planned for 60 articles, with the aim of achieving a maximum confidence interval of 20%. After completing collection for the 95 articles for our main analysis, we identified only one article that selected the non-default option. To save time, while still achieving a 20% confidence interval we decided to continue collecting data until we had 30 mixed-design ANOVA calculations, as opposed to 60. We miscounted how many articles in our dataset included mixed-design ANOVA and ended up coding 36 instead of 30. In the end, our confidence interval for selecting the non-default option (2-19%) is less than the originally planned 20%.
- We split study objective 3 from the protocol into study objectives 3 and 4 in this manuscript. The objectives themselves did not change. We feel it’s easier to interpret the results section of this manuscript when these objectives are presented separately.
- This manuscript presents our results in slightly different formats than what was proposed in the protocol. For example, some tables are split up and a few columns and rows differ. These decisions were made after analyzing the data and with the aim of presenting the results in a clear manner.
- When finalizing the writing of this manuscript, we realized there are two articles included in our results that were only coded by a single investigator (ids 143 and 146). For these two articles, one investigator selected that they did not have enough statistical expertise to code the article, but a second coding was not completed by another team member.
- The protocol did not put a limit date on our search query. A limit end date must be used to reproduce the query results.
- We made minor edits to the coding during data collection. These include:

- Changing the coding for the mixed-design ANOVAs to simply ask whether or not the default option was used, without making a judgment as to whether it was appropriate to use the default option. We made this change after coding approximately 50 articles and because we felt we could not always accurately state whether or not it was the appropriate choice.
- We added the option to code the type of power calculation as ‘solving for sample size after the study was complete’. We only coded this option when it was unambiguous that the sample size calculation was performed after the study was complete.
- For the question about errors in the sample size calculation, the coding form had the options “unsure” and “likely”. Before analyzing the data, we collapsed these options together because we found it difficult to be consistent within and among coders regarding these two coding options.

## Supplementary Material B. Additional tables and figures

**Supplementary Table 1.** Estimated number of articles published between 1 Jan 2017 and 31 May 2022 that reference G*Power.

|  | PubMed Central (95% CI) | PubMed (95% CI) |
| --- | --- | --- |
| Any power calculation | 21000 (20000 - 22000) | 48000 (46000 - 49000) |
| Sample size calculation | 14000 (13000 - 16000) | 32000 (28000 - 36000) |
| ANOVA sample size calculation | 3000 (2000 - 5000) | 8000 (5000 - 11000) |

We excluded articles that discuss G\*Power, but do not report using this software for a power calculation. The table includes rows for publications that report using G\*Power for any power calculation related to any statistical test (any power calculation), a power calculation for any statistical test that solves for sample size (sample size calculation), and a power calculation for an ANOVA that solves for sample size (ANOVA sample size calculation). The total number of articles in each database from 1 Jan 2017 to 31 May 2022 is: PubMed Central 3,285,893; PubMed 7,318,980. Numbers are rounded to the nearest thousand to avoid suggesting a higher level of precision than our method of estimation can provide.

**Supplementary Figure 1.**
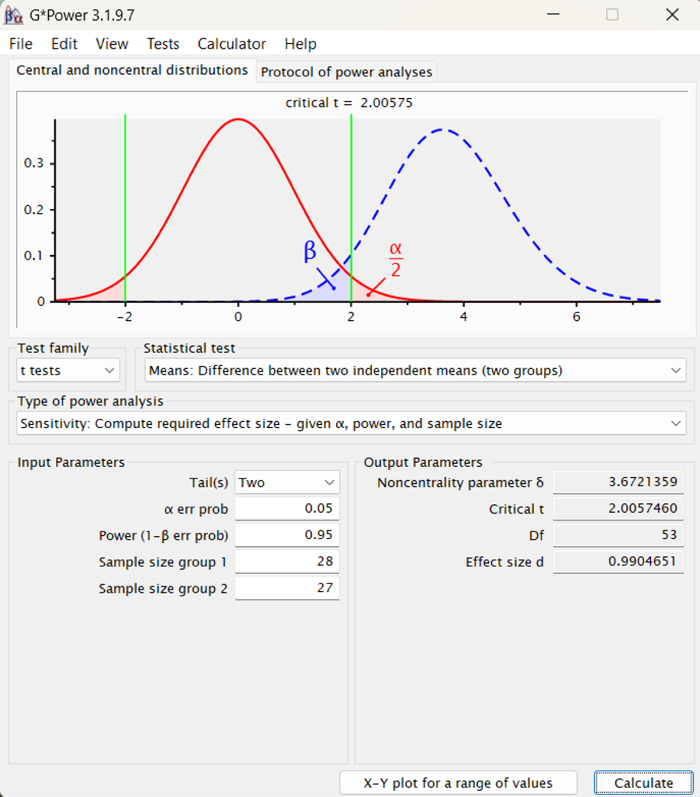
Sensitivity calculation for two-sample t-test with n = 55 and power = 95%.

**Supplementary Figure 2.**
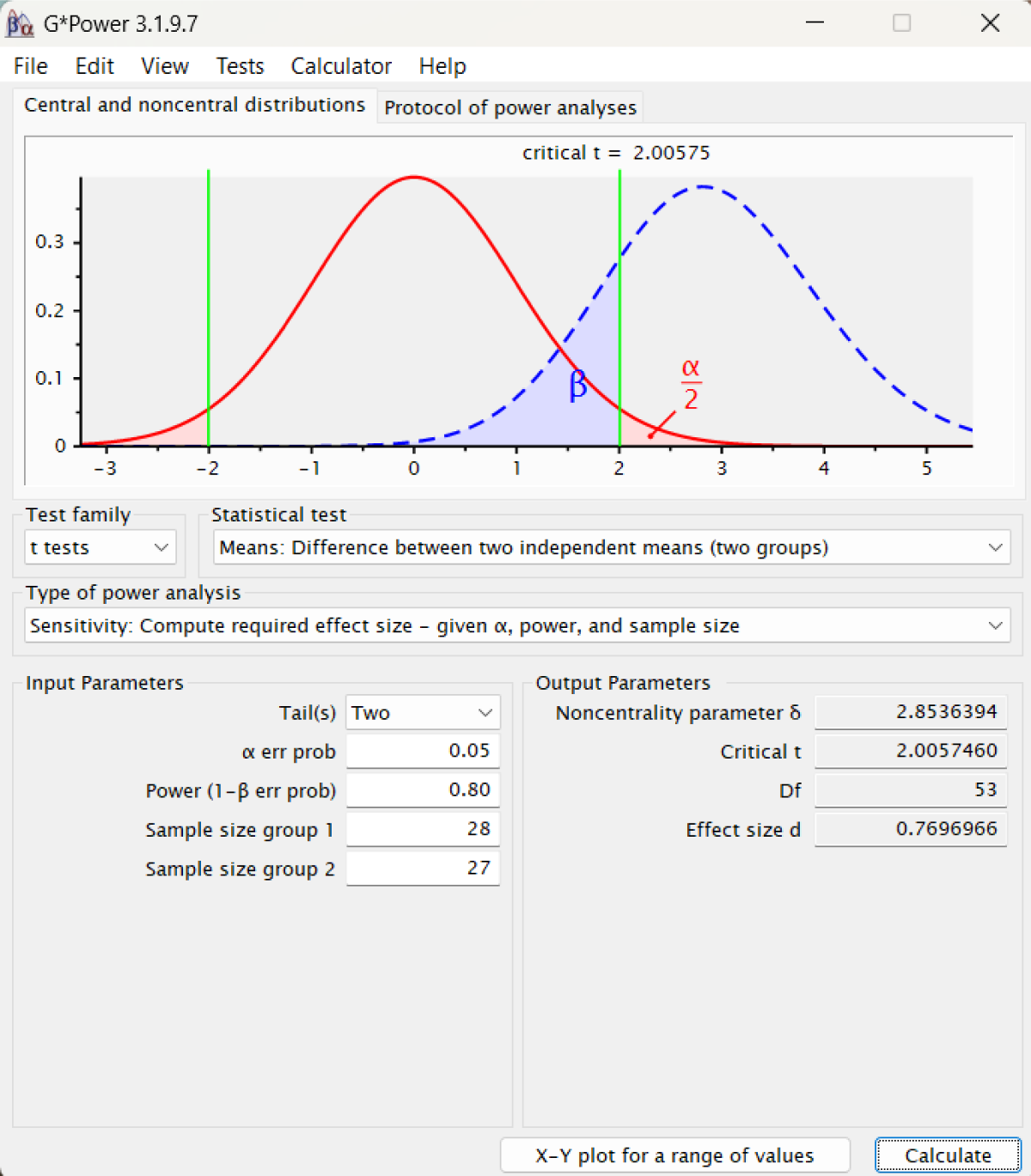
Sensitivity calculation for two-sample t-test with n = 55 and power = 80%.

## Supplementary Material C. Inter-rater agreement

**Supplementary Table 2.** Inter-rater agreement.

| Variable | Cohen's k | Percent agreed | n disagreed | n total | n categories |
| --- | --- | --- | --- | --- | --- |
| id | 1.00 | 100% | 0 | 147 | 147 |
| pmcid | 1.00 | 100% | 0 | 147 | 147 |
| protocol | 0.65 | 97% | 5 | 147 | 2 |
| include | 0.72 | 98% | 3 | 147 | 2 |
| participants | 0.82 | 96% | 5 | 141 | 2 |
| journal | 0.73 | 74% | 37 | 141 | 141 |
| pub_year | 0.95 | 96% | 6 | 141 | 7 |
| impact_factor | 0.82 | 83% | 24 | 141 | 112 |
| power_calc_type | 0.68 | 84% | 23 | 141 | 10 |
| multiple | 0.77 | 89% | 16 | 141 | 3 |
| version | 0.95 | 98% | 2 | 93 | 3 |
| version_text | 0.99 | 99% | 1 | 93 | 13 |
| power | 0.93 | 99% | 1 | 93 | 3 |
| power_text | 0.89 | 94% | 5 | 85 | 13 |
| alpha | 0.85 | 97% | 3 | 93 | 3 |
| alpha_text | 1.00 | 100% | 0 | 80 | 4 |
| sample_size | 0.34 | 92% | 7 | 93 | 3 |
| sample_size_text | 0.88 | 88% | 10 | 82 | 70 |
| effect_size_type | 0.84 | 89% | 10 | 93 | 10 |
| effect_size_value | 0.81 | 94% | 6 | 93 | 3 |
| effect_size_value_text | 0.95 | 95% | 3 | 65 | 34 |
| stat_test | 0.68 | 84% | 15 | 93 | 7 |
| reproducible | 0.23 | 70% | 28 | 93 | 4 |
| justification | 0.48 | 61% | 36 | 93 | 15 |
| just_previous | 0.43 | 82% | 17 | 93 | 2 |
| just_pilot | 0.48 | 96% | 4 | 93 | 2 |
| just_convention | 0.75 | 90% | 9 | 93 | 2 |
| just_mcid | 0.39 | 97% | 3 | 93 | 2 |
| just_none | 0.56 | 78% | 20 | 93 | 2 |
| just_ref | 0.21 | 88% | 11 | 93 | 2 |
| just_other | -0.02 | 96% | 4 | 93 | 2 |
| mult_compare | 0.24 | 67% | 31 | 93 | 4 |
| anova_within_between | 0.54 | 69% | 29 | 93 | 6 |
| match | 0.48 | 67% | 31 | 93 | 5 |
| error | 0.28 | 72% | 26 | 93 | 4 |

The variables are listed as they appear in the open data. See the data dictionary for a description of each variable. Cohen's kappa is mostly irrelevant for variables with a large number of categories, and can be ignored. Not all variables were relevant for all articles we coded; thus, 'n total' differs among the variables. 'justification' was coded as a multiple selection question with 7 options. We re-coded this variable into 7 binary variables and calculate the inter-rater agreement for each one.

- We calculated inter-rater agreement for all non-open-ended questions, as well as for open-ended questions for which a number was entered. For all these variables, Supplementary Table 2 shows Cohen’s kappa, percent agreed, number of disagreements, total number of articles coded for that variable, and the number of categories that were used for that variable.
- Five questions included “Unsure” as an option (power_calc_type, multiple, anova_within_between, match, and error). Not forcing coders to select an option likely reduced the inter-rater agreement.
- For two questions, multiple options could be selected. This led to many possible categories. For example, the 5 options for the multiple selection question for the power_calc_type variable leads to 32 possible combinations (2^5). For these questions, if coders agreed on some of the selections, but not all of them, then it was coded as a disagreement.
- Variables with text response (e.g., journal) may have small differences that were counted as disagreements (e.g., typos).
- Many of the variables have *n total* of 93 rather than 95 because two papers were not coded by a second investigator (see Supplementary Material A).
- Disagreements for the variable impact_factor mostly stem from (1) an issue one coder had, where he could not access impact factors, and (2) inputting the wrong year (our coding form asked to enter the impact factor from two years before the publication. This would have been the impact factor available at the time the authors of the sample size calculation were publishing their manuscript.
- Reproducibility had somewhat low agreement. This partially stemmed from one author’s greater familiarity with the G*Power software and ability to make assumptions. Although, perhaps it’s best to assume that someone reading an article would not be familiar with G*Power and thus not able to make these assumptions.

## Footnotes

1 As we produced estimates rather than tested hypotheses, we did not make adjustments to account for multiple comparisons. We aimed to balance the time it takes to code articles with the precision of our results, which need not be highly precise for the purposes of this study. For example, if we discover that 20-40% (95% CI) of power calculations are reproducible, we believe this remains a problem worth addressing regardless of whether the true value is 20% or 40%.

2 We made this assumption to help calculate a relevant sample size, it is not a hypothesis that we aimed to test.

3 For a nearly equivalent search today, use the query (GPower OR “G Power”) AND (“2017/01/01” [Publication Date] : “3000” [Publication Date]) AND (“1000” [pmclivedate] : “2022/05/31” [pmclivedate]). This query removes articles that PubMed Central indexed after our search date. The values “3000” and “1000” can be interpreted as “3000/12/31” and “1000/01/01”, they are upper and lower date limits, they are not special values.

4 We originally planned to code 60 articles for mixed-design ANOVAs. Deviations from our original protocol are detailed in Supplementary Material A.

5 Within our sampling time frame, PubMed indexed about 2.23 times more articles than PMC (7,318,980 articles in PubMed / 3,285,893 articles in PMC.). If we assume that articles indexed in PubMed report using G*Power at a similar rate as articles indexed in PMC, and that all articles indexed in PMC are also indexed in PubMed, then we can multiply 21,000 by 2.23 and arrive at about 48,000 (Supplementary Table 1).

6 See variable *reproducible_text* from the data for more detail about the assumptions we made.

7 We use the term *likely* because—without additional information—we can’t be certain we reproduced these calculations. It is possible to arrive at the same sample size with different combinations of parameters. We made assumption based on our experience with sample size calculations and G*Power.

8 Due to calculating the point estimates using Monte Carlo simulation, some numbers may appear to be rounded in the wrong direction. For example, this manuscript rounds 66/95 to 70%, even though 66 divided by 95 is 69.47%. The Monte Carlo simulation estimated a number just over 69.5%, which we then rounded to 70%.

9 We only coded the statistical test as reported if it was identified alongside the sample size calculation. While one could argue that the statistical test can be inferred by the effect size type, or by looking at the results sections, this proved difficult. If details about the statistical test were missing, but the general statistical test was mentioned, we coded the statistical test as reported. There were several cases where all 6 elements were reported, but we could not reproduce the calculation.

10 Publisher names were identified using Clarivate’s Journal Citation Reports and coded according to the publisher names used on their website.

11 This finding is in large part because 66% of the sample size calculations did not explicitly state what statistical test they were using. In some cases, we could assume what test was used based on effect size types and other contextual information.

12 Details available in the *impact* column of the open data.

13 We first heard this term from a twitter post by Dorothy Bishop: https://twitter.com/deevybee/status/1628736447665635328

14 See Goodman and Berlin (1994) for an explanation of why *post-hoc* power calculations are problematic.

15 In some disciplines, sample sizes that are too big are seen as wasteful and can be a bigger issue (sacrificing animals, exposing patients to treatments with unknown side effects). We focus on what we consider the more widespread issue of sample sizes that are too small..

16 We performed these calculations based on data from the 2015–2018 National Health and Nutrition Examination Survey (NHANES), which include data from more than 10,000 people in the United States. Notably, the effect size is larger when analyzing more uniform subsets of the population (e.g., for ages 20-29: *d* = 1.46; for non-Hispanic white people of age 20-39: *d* = 1.69). Calculations are provided in the analysis script.

17 For example, “We performed a power calculation using G*Power 3.1.9.7 for a two-tailed independent sample t-test with alpha = .0167 (Bonferonni corrected for 3 comparisons), and power = .80. Powering for a medium effect size of Cohen’s *d* = 0.5, requires a sample size of 64 participants per group”.

